# HIPTox – Hazard Identification Platform to Assess the Health Impacts from Indoor and Outdoor Air Pollutant Exposures, through Mechanistic Toxicology: A single-centre double-blind human exposure trial protocol

**DOI:** 10.1101/2023.12.13.23299801

**Authors:** Thomas Faherty, Huda Badri, Dawei Hu, Aristeidis Voliotis, Franics D. Pope, Ian Mudway, Jacky Smith, Gordon McFiggans

**Author notes:** These authors contributed equally to this work (Thomas Faherty, Huda Badri).

## Abstract

Over the past decade, our understanding of the impact of air pollution on short- and long-term population health has advanced considerably, focusing on adverse effects on cardiovascular and respiratory systems. There is, however, increasing evidence that air pollution exposures affect cognitive function, particularly in susceptible groups. Our study seeks to assess and hazard rank the cognitive effects of prevalent indoor and outdoor pollutants through a single-center investigation on the cognitive functioning of healthy human volunteers aged 50 and above, with a familial predisposition to dementia. Participants will all undertake five sequential controlled exposures. The sources of the air pollution exposures are wood smoke, diesel exhaust, cleaning products, and cooking emissions, with clean air serving as the control. Pre- and post-exposure spirometry, nasal lavage, blood sampling, and cognitive assessments will be performed. Repeated testing pre- and post-exposure to controlled levels of pollutants will allow for the identification of acute changes in functioning as well as the detection of peripheral markers of neuroinflammation and neuronal toxicity. This comprehensive approach enables the identification of the most hazardous components in indoor and outdoor air pollutants and further understanding of the pathways contributing to neurodegenerative diseases. The results of this project have the potential to facilitate greater refinement in policy, emphasizing health-relevant pollutants and providing details to aid mitigation against pollutant-associated health risks.

## 1. Introduction

### 1.1. Background and Rationale

Air pollution is the primary global environmental risk to health that increases mortality worldwide [1]. Adverse effects of low-quality air on cardiovascular and respiratory systems are well-established [2,3], with our understanding of its effects on both short- and long-term population health advancing considerably over the last decade. There is also growing evidence that air pollution is neurotoxic [4,5], degrading the brain’s cellular structures [6], leading to neurocognitive decline [7] and delaying neurocognitive development [8]. However, there are still major gaps in our understanding of the most harmful components within the air we breathe and the mechanisms by which they induce adverse effects [9].

Lifetime exposure to low-quality air has been linked to greater risk of substantial cognitive deficits associated with progressive neurodegenerative diseases such as Alzheimer’s Disease [10], Parkinson’s Disease [11], and Multiple Sclerosis [12]. Chronic exposure to low-quality air is also associated with poorer than expected memory [13–16], psychomotor and sensory processing [17], and cognitive executive function [18,19] in clinically healthy children. Short-term episodes of exposure to low-quality air have also shown impact on these cognitive functions [20–23]. Evidence is, however, mixed with regards to the impact of certain pollutant mixtures on different cognitive domains [7]. It is therefore critical to assess a range of cognitive functions and changes as a function of differing common sources of air pollution.

Air pollution may manifest its acute and chronic impacts on the brain via direct interaction of inhaled particles, or desorbed chemical constituents, with non-neuronal glial cells and neurons in the brain, based either on their uptake via translocation along olfactory neurons to the olfactory bulb [24]; or their entry across the blood brain barrier from the circulation [25]. This has been supported by evidence of combustion-like nanoparticles in the brains of sentinel animals and humans from polluted environments [26], associated in autopsy samples with histopathological features of early dementia, microglia immune defence activation and neuronal injury [27]. A second hypothesis proposes that the neurological damage reflects an indirect impact of air pollution induced systemic inflammation and its transmission to the brain across the glial-neurovascular unit [28,29]. There is therefore a clear necessity for this study to investigate evidence for both routes through both collection of blood to measure traditional toxicity endpoints, including pro-inflammatory biomarkers and markers for organ specific toxicity, and nasal lining fluids to identify the presence of neuronal injury markers.

We believe there is a strong need to respond to the emerging evidence of the impact of air pollution on the brain across the life course, from early life cognitive delays [14], adverse mental health [30] to dementia risk. Moreover, the estimated cost of air pollution is £20 billion per year in the UK, specifically attributable to cardiopulmonary mortality and morbidity alone [31]. The neurological effects carry significant economic and societal risks, above and beyond the cardiopulmonary risks, and will likely increase the economic and societal cost significantly. An enhanced understanding of which components of the air are drivers of these adverse neurological effects and the underlying causal mechanisms are therefore questions of fundamental importance to the health and economics of the nation.

This presented single-centre, double-blind, human exposure trial will investigate the effect of exposure to four common air pollutants deriving from: cleaning products; diesel exhaust; cooking emissions; and woodsmoke, on cognitive function comparative to clear air. Coupled to complementary cell and transgenic animal model exposures in the same programme, the data generated from these human exposures will allow the identification of the most hazardous components of outdoor and indoor air pollutants and to elucidate the causal pathways contributing to neurodegenerative disease development and exacerbation. The results of this project will facilitate greater refinement in policy to place greater emphasis on health-relevant pollutants and provide these details to aid mitigation against pollutant associated health risks.

### 1.2. Objectives

The primary objective of this study is to rank the acute effects of a range of air pollution exposures on cognitive function in healthy aged individuals with a family history of increased dementia risk.

The secondary objective of this study is to identify evidence of changes in inflammatory or immune signals post exposure to pollutants compared to clean air, exploring possible underlying mechanisms of any identified cognitive changes.

### 1.3. Trial Design

This is a single centre double-blind study of the effects of mixed air pollutants on the cognitive function of healthy human volunteers with a family history of increased risk of dementia. Participants will all undertake sequential controlled exposures to four air pollutants and clean air as control exposure, i.e., a repeated measures design. Cognitive testing, biological sampling and physiological tests will be carried out before and after each exposure. The study design is outlined in Figure 1.

**Figure 1.**
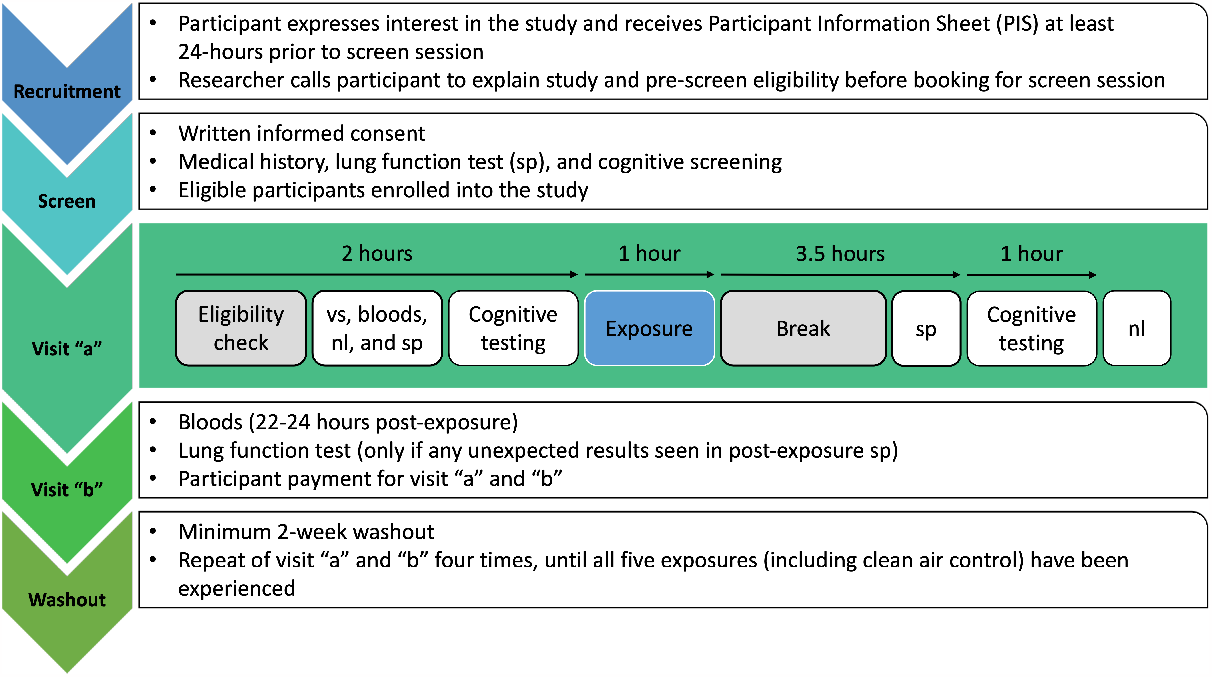
Study data collection procedure. vs = vital signs; nl = nasal lavage; sp = spirometry (lung function)

## 2. Materials and Methods

### 2.1. Study Setting

The HIPTox study will be carried out at the NIHR/Wellcome Trust Clinical Research Facility (MCRF), Grafton Street, Manchester and the Manchester Aerosol Chamber (MAC), Simon Building, University of Manchester, Manchester, UK.

### 2.2. Participant volunteers

This study will aim to recruit 45 clinically healthy individuals aged 50 and above with a family history of dementia. See Appendix A for full inclusion and exclusion criteria. Recruitment began in May 2023 and due to significant challenges, as explained in detail in the discussion section, the originally planned sample size was reduced to 15 participants. The study is currently ongoing, and data collection is expected to be completed by the end of 2023. See Table 1 for study timeline.

**Table 1.** Study time schedule of enrolment, exposures, and visits. Exp = Exposure.

|  |  | Recruitment | Enrolment |  | Post-enrolment |  |  |  |  |  |  |
| --- | --- | --- | --- | --- | --- | --- | --- | --- | --- | --- | --- |
|  |  |  | First Wave (W1) | Second Wave (W2) | Exp 1 (W1) | Exp 2 (All) | Exp 3 (All) | Exp 1 (W2) | Exp 4 (All) | Exp 5 (All) | Remnant Exp(s) |
|  | Start date | 09-Jan-23 | 02-May-23 | 01-Jun-23 | 15-May-23 | 19-Jun-23 | 24-Jul-23 | 21-Aug-23 | 11-Sep-23 | 16-Oct-23 | 20-Nov-23 |
|  | End date | 30-Jun-23 | 31-May-23 | 30-Jun-23 | 02-Jun-23 | 07-Jul-23 | 11-Aug-23 | 25-Aug-23 | 29-Sep-23 | 03-Nov-23 | 30-Nov-23 |
| ENROLMENT |  |  |  |  |  |  |  |  |  |  |  |
|  | Information Sheet | X |  |  |  |  |  |  |  |  |  |
|  | Informed Consent |  | X | X |  |  |  |  |  |  |  |
|  | Eligibility Screen |  | X | X | X | X | X | X | X | X | X |
| VISITS |  |  |  |  |  |  |  |  |  |  |  |
| Exp 1 | Visit A |  |  |  | X |  |  | X |  |  | X |
|  | Visit B |  |  |  | X |  |  | X |  |  | X |
| Exp 2 | Visit A |  |  |  |  | X |  |  |  |  | X |
|  | Visit B |  |  |  |  | X |  |  |  |  | X |
| Exp 3 | Visit A |  |  |  |  |  | X |  |  |  | X |
|  | Visit B |  |  |  |  |  | X |  |  |  | X |
| Exp 4 | Visit A |  |  |  |  |  |  |  | X |  | X |
|  | Visit B |  |  |  |  |  |  |  | X |  | X |
| Exp 5 | Visit A |  |  |  |  |  |  |  |  | X | X |
|  | Visit B |  |  |  |  |  |  |  |  | X | X |
| MEASURES |  |  |  |  |  |  |  |  |  |  |  |
|  | Pre-Exposure |  |  |  | X | X | X | X | X | X | X |
|  | Post-Exposure |  |  |  | X | X | X | X | X | X | X |

#### 2.2.1. Power Calculation

Based on effect size and variance from a previous unpublished study looking at cognitive load and attention (measured as change in response time (ΔRT) after Diesel Exhaust exposure (ΔRT = 22ms, s.d. = 25ms), a sample size of 30 subjects will have 90% power to detect significant response time changes of 15ms assuming the standard significance level of p<0.05. We acknowledge this is a different design utilising an older adult population and have therefore calculated for a slightly smaller effect size than that of the previous study to compensate for larger RT variance in older adults. As longitudinal studies are prone to extensive participant attrition, the sample size of 45 aims to allow full data to be collected for a minimum of 30 participants.

In response to recruitment challenges, we revised the sample size, prompting a re-exploration of the sample size estimate and power calculation to identify the minimum sample likely to yield valuable results. The proposed study design involves comparing the impact of four pollution exposures by examining the differences between pre- and post-exposure measurements relative to baseline measurements. The independent design from the previous study, featuring one measure per group, makes it unfeasible to extrapolate data. Therefore, under the assumption of a conservative correlation among repeated measures set at 0.5, a minimum sample size of 11 participants is needed to achieve 90% power in detecting a moderate (0.45) effect size at a significance level of 0.05.

#### 2.2.2. Patient and Public Involvement and Engagement (PPIE)

This project was supported by the Manchester University Foundation Trust’s public and patient engagement team (VOCALS) during the study conception and funding approval. Numerous public and patient engagement activities were carried out prior to protocol finalisation. These events helped shape the final study design including duration of the study. During the recruitment phase we continued to engage with patients and the public via over 45 engagement events, including “Brain Health Day, Manchester”, local workshops and pollution awareness events. We aim to carry out a post study completion engagement event with all the participants to disseminate the results alongside publication in scientific journals.

### 2.2.3. Recruitment

Participants will be recruited from the general public. Potential volunteers who are employees at The University of Manchester or Manchester University NHS Foundation Trust may take part if not involved with the project. Advertisement posters will be placed in any public spaces including trust and university buildings with contact details of the research team on; interested participants will be able to contact for further details. Social media advertisement using official accounts of the University of Manchester, NHS Manchester Foundation Trust, dementia charities, local community groups, and places of worship. We will also contact participants who have previously given permission to be contacted for research studies.

To promote participant retention, participant payment will increase each visit so that there is an increasing incentive to continue participation as the study continues. Informal participant feedback will be collected each visit to improve subsequent visit comfort and amend the protocol as necessary. Data from those who discontinue participation, whether through personal withdrawal or change in eligibility, will be included in data analysis where inclusion of partial data is possible.

#### 2.2.4. Participant blinding

This will be a double-blind study with both the trial participants and researchers / care staff responsible for data collection blinded to the exposure. Those responsible for data collection are MCRF staff, Dr Thomas Faherty, and Dr Huda Badri. Other investigators will not be blinded as to the order of exposures. Participants are aware of what the 5 pollution exposures are, but not the order in which they will experience them. Participants will be asked by the (unblinded) MAC team which of the 5 exposures, including clean air, they believed they experienced following each exposure as well as a confidence judgement. Trial participants and data collection staff will also be blinded during data analysis.

The PI will make the decision to un-blind the current participant exposure in the case of a serious adverse event (SAE) when knowledge of the chemical composition of the exposure is vital for appropriate clinical management or the participant’s well-being.

### 2.2.5. Participant safety and anonymity

All participants enrolled in this study will be allocated a unique anonymous identifying code which will appear on all data collection documents. The hard-copy case report forms (hCRF) and source data files (SDF) will be kept within a locked room. All electronic data will be stored on secure password-protected servers with limited access to members of the research team.

Participant eligibility will be reviewed at the beginning of each visit (separated by a minimum two-week washout period) prior to air pollution exposure. Participation will be discontinued for those who do not match the eligibility criteria when reviewed.

The checking for the occurrence of adverse events (AEs) will begin from enrolment and will continue for the individual participant until their last visit. At each study visit, the researcher will assess eligibility and recent clinical history including the occurrence of adverse or serious adverse events (SAEs). Details of adverse and clinical events will be captured on the trial hCRF and eCRF. All SAEs will be recorded in the hospital notes, the eCRF, the hCRF, and the Sponsor’s SAE Recording Log. All SAEs will be reported to the Sponsor via the Research Office (RO) dedicated mailbox on an SAE form and the SAE Log will be sent to Sponsor on request. The PI will complete the Sponsor’s SAE form and the form will be sent within 24-hours of the Investigator becoming aware of the event. The PI will respond to any SAE queries raised by the Sponsor as soon as possible.

Expected Adverse Reactions are possible short-term respiratory symptoms such as cough/ shortness of breath to susceptible individuals. We will minimise this risk by assessing the participants pre-enrolment onto the study for respiratory conditions. We will also conduct safety measures such as spirometry and exclude individuals with abnormal results which may also suggest a high risk to pollutant exposure. Investigators will complete the appropriate SAE form on the eCRF and an automatic email notification will be sent to the Trial Manager, PI and Sponsor.

A Suspected Unexpected Serious Adverse Reaction (SUSAR) is an adverse reaction that is classed as both serious and unexpected. The Trial Manager will ensure that SUSAR reports are un-blinded and reviewed by the PI or designee within 2 days and adjudicate whether the event constitutes a SUSAR. The Trial Manager will ensure that fatal or life-threatening SUSARs are reported to UREC as soon as possible, but no later than 7 calendar days after the receipt of the eSAE report. Any additional information will be reported within 8 days of sending the first report.

### 2.3. Exposures

The four chosen pollutants have been selected as comparators to clean air as they are known to contribute substantial fractions to indoor and outdoor air.

#### Diesel exhaust

Previous studies demonstrate a clear negative effect on cardiopulmonary health [32,33] and there is evidence of neurological impacts of diesel exhaust in young healthy volunteers [34,35], therefore a pollutant of interest in this study.

#### Woodsmoke

Woodsmoke is a prevalent indoor and outdoor air quality issue in the UK [36]: in the last 10 years its emissions have substantially offset the decrease in particulate matter (PM) from other sources, increasing by 33%, and it is likely to continue increasing in the medium-term [37], therefore identifying the impact on human cognitive health is of necessity at this critical juncture.

#### Cooking emissions

The significant contribution of cooking aerosol to indoor/urban air quality has recently emerged as an important topic within the aerosol community [38], but there is little solid evidence on its potential health impacts. A recent study did suggest that ultrafine aerosols from frying meat resulted in altered electrical brain activity in human subjects [39], warranting its further investigation here.

#### Cleaning products

Minimal evidence exists linking secondary organic aerosol (SOA) formed from household cleaning products to adverse effects on cognitive function despite its abundance in indoor air.

Participant exposures will be conducted at the Manchester Aerosol Chamber (MAC) facility. The MAC facility is part of the ATMO-ACCESS atmospheric simulation chamber network. It is an 18m3 Fluorinated ethylene propylene Teflon bag housed in a temperature and humidity-controlled enclosure and illuminated by a combination of wavelength filtered arc and halogen lamps. Clean inlet air is ensured by scrubbing with a series of filters and conditioners. Particle and gas levels are controlled either by injection of individual components, or by coupling to a variety of real pollutant sources. Please see the Centre for Atmospheric Science webpage [40] and the following paper [41] for comprehensive details. This facility as well as the techniques and specific instruments employed here have been used in many previous chamber studies [42–52]. For each pollutant mixture, a full characterisation will be conducted to identify the exact chamber conditions and experimental setpoints to achieve target pollutant concentrations to within acceptable limit. All experimental setpoints are computer-controlled and continuously monitored, ensuring participant safety, and enabling repeatability.

Participants will undertake a 60-minute exposure to a controlled concentration of common air pollutants in addition to a clean air exposure to act as a control. Participants will be fitted with a mask covering the nose and mouth (VariFit™ NIV non vented mask) attached to the aerosol chamber via a plastic pipe. The mask will use a two-way valve (Anti-pollution T-piece directional valve) allowing participants to inhale air from the chamber without forced air pressure and exhale into the surrounding room, avoiding contamination of the chamber air mixture. See Figure 2.

**Figure 2.**
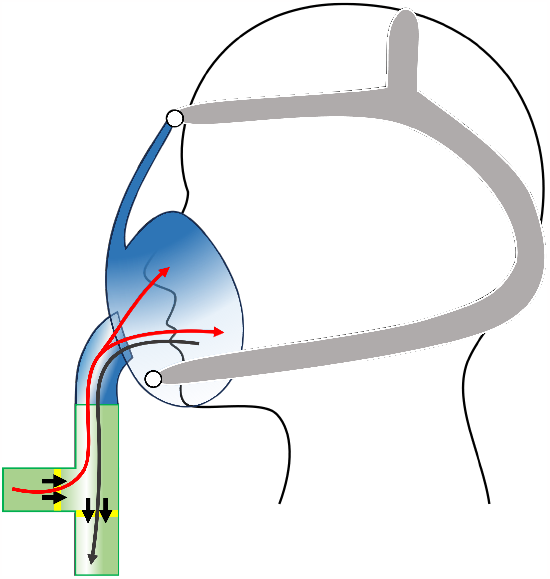
The two-way valve and mask set-up during exposure. Black arrows show airflow direction of one-way valves. Participants inhale air from the chamber (red line) and exhale into the surrounding room (dark grey line).

A review of adverse event reporting from previous woodsmoke and diesel exhaust exposure chamber studies utilising higher pollutant concentrations showed no evidence of adverse events. See supplementary file 2 for full details.

### 2.4. Physiological measures

#### 2.4.1. Spirometry

Spirometry will be a safety measure in this study. Volunteers will be asked to perform spirometry, to achieve 3 reproducible Forced expiratory volume (FEV1) and forced vital capacity (FVC) measurements according to the American Thoracic Society/European Respiratory Society guidelines[53].

#### 2.4.2. Blood tests

Blood will be taken as part of the assessment of inflammatory response. This will be plasma and RNA samples 5mls (3 bottles pre-exposure and 3 bottles on visit b), a total of 30mls per participant per visit.

These collected samples will be centrifuged and transported to Imperial College Health Care NHS trust for analysis and storage. These will be stored for 2 years under their Human Tissue Authority License number 12275.

Once analysis has been completed, any additional samples will be stored in biobanks for up to 30 years if participants have consented to this optional/additional section in the ICF. If a participant does not wish to have their samples stored in a biobank, this will not affect their participation in the study. It is expected that these biobanked samples will be accessible to researchers for further analysis in the future.

#### 2.4.3. Nasal lavage

Participants will undertake nasal lavage with 15ml pre-warmed normal saline solution (0.9% NaCl). Sterile solution will be instilled into the participant’s nose whilst their head is flexed and chin towards their chest. 10 sprays in one nostril, whilst the other is closed off. This will be repeated four times and then using a glass beaker the fluid is collected. The whole process is then repeated with the other nostril; a total of 50 sprays in each nostril. The sample is then decanted into a centrifuge tube and centrifuged for 10mins at 800g at 4°C.

The nasal lavage samples will then be transported to Imperial College Health Care NHS trust for analysis and storage. These will be stored for 2 years under their Human Tissue Authority License number 12275. These samples will then be destroyed as per the terms of the license.

### 2.5. Cognitive measures

#### 2.5.1. Dementia assessment

The General Practitioner Assessment of Cognition[54] (GPCog) is a screening tool for global cognitive impairment seen in those with dementia. The Participant Examination assessment (See supplementary file 3) will be completed as part of eligibility screening. Briefly, participants are given a name and address to immediately repeat and remember. Participants are then asked to provide the exact day, complete two diagrams highlighting clock time, and tell the experimenter something that has happened in the news in the past week. Participants then recall the name and address they were asked to remember at the start. Usually, a score of 8 or below would involve additional information (Step 2: Informant Interview) from an informant, i.e., family member who would know the participant and their current functioning compared to previous years. As we will not have access to an informant, participants will be allowed one error, so a score of 8 or 9 will indicate that participants are cognitively intact and can continue with the study.

#### 2.5.2. Cognitive tasks

For cognitive assessment, we will utilise five tasks — one standardised manual task and four internally designed computer tasks. Two of the computer tasks will integrate novel elements into previously used paradigms, while two will adhere to more commonly used methodologies, ensuring alignment with established research. This approach combines standardised rigour with innovative, internally developed tasks, providing a comprehensive evaluation of cognitive domains. A summary of the five tests is detailed in Table 2 and further description is below. Full methodology of each task is provided in supplementary materials (file 4).

**Table 2.**
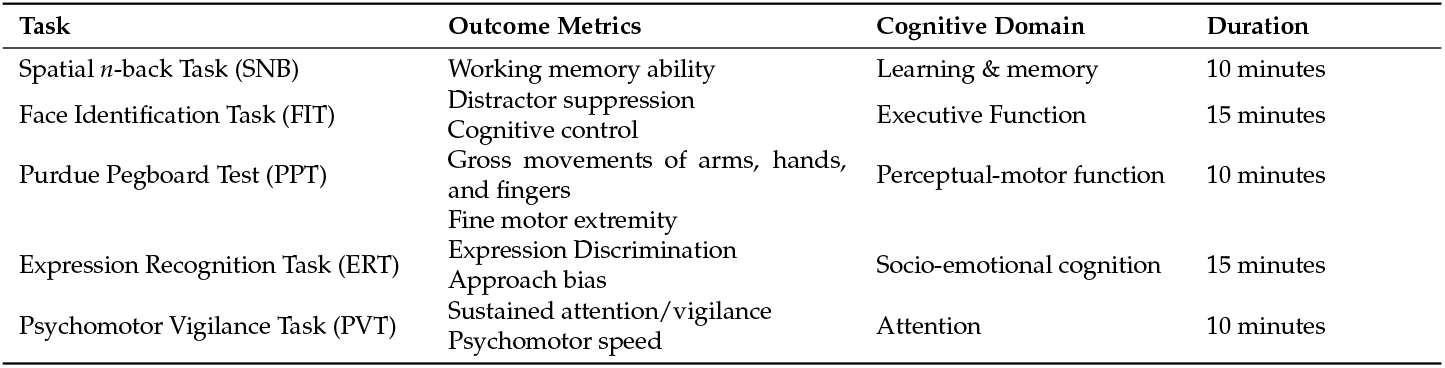
Summary of the cognitive tasks completed by participants pre- and post-exposure. Tasks will be administered in table order, starting with the Spatial n-back Task (SNB).

| Task | Outcome Metrics | Cognitive Domain | Duration |
| --- | --- | --- | --- |
| Spatial *n*-back Task (SNB) | Working memory ability | Learning & memory | 10 minutes |
| Face Identification Task (FIT) | Distractor suppression<br>Cognitive control | Executive Function | 15 minutes |
| Purdue Pegboard Test (PPT) | Gross movements of arms, hands, and fingers<br>Fine motor extremity | Perceptual-motor function | 10 minutes |
| Expression Recognition Task (ERT) | Expression Discrimination<br>Approach bias | Socio-emotional cognition | 15 minutes |
| Psychomotor Vigilance Task (PVT) | Sustained attention/vigilance<br>Psychomotor speed | Attention | 10 minutes |

The Purdue Pegboard Test[55] (PPT) is a manual test not requiring a computer. The other four cognitive tasks will require a Windows 10 computer running Matrix Laboratory (MATLAB) version R2022a [56]. These four tasks are in the format of a MATLAB script utilising the Psychophysics Toolbox version 3.0.18[57].

##### Spatial *n*-Back Task

A working memory task using spatial locations of stimuli. This is a measure of participant ability to hold multiple pieces of information in the brain at one time and recall that information when necessary.

##### Face Identification Task

A selective attention task using particularly distracting face stimuli. This is a measure of cognitive control, a facet of executive function, specifically participant ability to ignore distracting stimuli and focus on task goals.

##### Purdue Pegboard Test

This is a standardised test to measure of motor control, specifically participant hand and finger dexterity to manoeuvre small objects.

##### Expression Recognition Task

A Go/No-Go task using affective facial stimuli. This is to measure socio-emotional cognition, specifically participant ability to identify the emotion expression of a human face at speed.

##### Psychomotor Vigilance Task

A simple reaction time task. This measures both sustained attention, i.e., participant ability to focus their attention for a sustained period of time, and psychomotor speed, i.e., speed of their response to changes in the visual field.

### 2.6. Data analysis

Data generated by this study will be analysed using longitudinal statistical modelling designed to account for the repeat measures in individual subjects (e.g., general estimating equation or random effects modelling, depending on the extent and nature of any missing or partial data). This will describe and account for changes in pre-exposure measures over time, comparing the effects of the different air pollution exposures to the clean air control.

Cognitive subscales will be entered into this process separately, to aid in pairing aerosols with undesirable changes identified in each cognitive domain.

In addition to ranking cognitive impacts in relation to the exposure sources, these data will be related to biomarkers of systemic and neuro-inflammation / injury to test the secondary hypothesis that acute cognitive effects and more severe downstream neurological impacts reflect underlying inflammatory processes.

Specifically, our primary and secondary end points are as follows:

#### Primary end point

Evidence of impairment in cognitive function post exposure as measured by any of these cognitive tasks compared to clean air:

1. Difference in the Approach bias measured via the Expression Recognition Task post pollutant exposure.
2. Change in the Cognitive Control response time post pollutant exposure.
3. Change in the 2-back discrimination index post pollutant exposure.

#### Secondary end point

1. Change in performance on any of these cognitive tasks post pollutant exposure compared to clean air:
  - Psychomotor Vigilance Task
  - Purdue Pegboard Test
2. Evidence of inflammatory or immune signals post exposure to pollutants compared to clean air.
3. Change in lung function post pollutant exposure compared to clean air.
4. Change in DNA post pollutant exposure compared to clean air.

### 2.7. Data management and monitoring

The study will be subject to the audit and monitoring regime of the University of Manchester. All research staff involved in this study will be familiar with the protocol and suitably qualified, trained, and competent to carry out the required techniques. Staff are also trained in ICH-GCP, local information governance policies and the data protection act. The source documents will be defined as the printed reports for any study measurement completed on a piece of equipment with this capability, or the case report forms designed by and completed by research staff. All research related documentation including the site file, case report forms etc. will be kept with the PI until the results of the study have been analysed and published. Then it will be boxed and archived offsite for a minimum of 5 years. Archiving and destruction logs will be generated.

## 3. Discussion

Using an integration of cognitive psychology, atmospheric sciences, clinical experimental medicine, and clinical biochemistry, this project aims to assess the comparative impact of source-specific aerosols on physiological and cognitive functioning. The methodology involves repeated testing pre- and post-exposure to controlled levels of pollutants, enabling the identification of acute changes in functioning and the detection of peripheral markers of neuroinflammation and neuronal injury.

We initially aimed for 45 participants; however, various recruitment challenges emerged during the study, such as the extended study duration, time-intensive visits, and the eligibility of those in the specific cohort of participants required. Despite efforts to alleviate the burden on participants through protocol reviews and an extensive recruitment drive involving over 45 Patient and Public Involvement and Engagement (PPIE) interactions, it became apparent that recruiting 45 participants was not feasible.

Upon reassessment, the study team adjusted the target to a smaller sample size of 15, assuming a similar dropout rate (33%). This adjustment was made to still maintain sufficient data for reasonable statistical power. Despite this adaptation, we acknowledge that the reduction in statistical power increases the likelihood of not detecting a true effect, leading to a higher risk of committing a Type II error due to wider confidence intervals. Detecting small effects is also challenging due to queries associated with using a shorter exposure time and concentrations relative to previous studies. Similarly, a smaller sample size is more susceptible to random fluctuations, which are anticipated in cognitive functioning. However, the repeated-measures design and control of participant activities during data collection aim to reduce this as a confounding variable.

We believe that, despite the increased risk of errors, the study is worthwhile with this smaller sample size, especially considering the feasibility of recruiting clinically healthy participants over the age of 50 with a family history of dementia. The anticipated results of this project hold the potential to inform policy refinement, placing greater emphasis on health-relevant pollutants. This detailed information will contribute to efforts aimed at mitigating health risks associated with exposure to pollutants.

## 4. Administrative Information

### 4.1. Reporting standards

The SPIRIT reporting guidelines were used to prepare this manuscript[58].

### 4.2. Trial registration

Trial registered on ISRCTN registration number 85634746, accepted on 3rd April 2023.

### 4.3. Protocol version

Current protocol version 1.6, 23rd February 2023.

### 4.4. Sponsor information

Sponsor details:

The University of Manchester

Ms Lynne Macrae, Faculty Research Practice Governance Coordinator

Faculty of Biology, Medicine and Health, 5.012 Carys Bannister Building, University of Manchester, M13 9PL

Telephone: 0161 275 5436

Roles and responsibilities of the sponsor:

The study sponsor has overseen the design of the study and will have oversight of the trial. The sponsor has ensured that the trial protocol, Participant Information Sheet (PIS), Informed Consent Form (ICF), General Practitioner (GP) letter, and submitted supporting documents have been approved by the University of Manchester Research Ethics Committee (UREC) and the Health Research Authority (HRA), prior to any participant recruitment taking place. This study will be conducted in compliance with the protocol approved by UREC and according to good clinical practice (GCP) standards and UK Clinical Trials Regulation.

The trial protocol, PIS, ICF, GP letter, and submitted supporting documents were approved by the HRA and UREC, before participant recruitment. All subsequent substantial protocol amendments will be documented and submitted for ethical and regulatory approval prior to implementation. Site-Specific Assessment from the Trust Research & Development (R&D) and NHS R&D approval were granted prior to participant enrolment.

### 4.5. Trial Status

The current protocol attached is version 1.6, dated 23rd February 2023. Recruitment began in May 2023 and the study is currently ongoing. It is expected that data analysis will begin in January 2024.

### 4.6. Dissemination

Our expectation is that after data analysis, information from this study will be widely disseminated in the medical and scientific community.

## Supporting information

Supplementary File 1 - Informed Consent Form

Supplementary File 2 - GPCog

Supplementary File 3 - Previous Chamber Study Safety

Supplementary File 4 - Cognitive Task Methodology

Supplementary File 5 - SPIRIT Checklist

## Author Contributions

All authors meet the ICMJE criteria for authorship. G.M. curated the work packages across the consortium. All authors were responsible for the study conception and design. J.S., T.F., and I.M. have overseen the development of the statistical analysis. H.B. & T.F. wrote the manuscript. All authors provided final approval of the manuscript.

## Institutional Review Board Statement

Ethical approval has been obtained from both the Health Research Authority (HRA) Integrated Research Application System (IRAS), ID 314890, and University of Manchester Research Ethics Committee (UREC), ID 2022-14762-25491.

## Informed Consent Statement

Informed consent, see supplementary file 1 for the informed consent form (ICF), will be obtained by the Principal Investigator (PI) and/or a nominated deputy as recorded on the sponsor’s Delegation of Responsibilities Log. All individuals taking informed consent will have received training in good clinical practice (GCP).

All interested participants will receive the participant information sheet (PIS) at least 24-hours prior to attendance for eligibility screening. Participants will be provided with a repeated full explanation of what is involved with the study when arriving for the eligibility screening session and given the opportunity to have any queries answered prior to providing written informed consent. Therefore, consent to enter this study will be obtained after a full account has been provided of its nature, purpose, risks, burdens and potential benefits, and the prospective participant has had the opportunity to deliberate.

## Data Availability Statement

There are currently no plans to share participant-level datasets.

## Acknowledgments

The authors would like to acknowledge and thank the following individuals and groups for their roles in successful study delivery:

- All volunteer participants.
- The Manchester Clinical Research Facility.
- The National Institute for Health and Care Research Manchester Biomedical Research Centre.
- The University of Manchester sponsorship team.
- Colleagues across all work packages in the HIPTox consortium

## Abbreviations

The following abbreviations are used in this manuscript:

eCRF: electronic case report form
ERT: Expression Recognition Task
FIT: Face Identification Task
GCP: good clinical practice
GPCog: general practitioner assessment of cognition
hCRF: hard-copy case report form
HRA: Health Research Authority
ICF: informed consent form
IRAS: Integrated Research Application System
MAC: Manchester Aerosol Chamber
MCRF: Manchester Clinical Research Facility
nl: nasal lavage
PI: principal investigator
PIS: participant information sheet
PM: particulate matter less than 2.5 micrometres in diameter
PPIE: Patient and Public Involvement and Engagement
PPT: Purdue Pegboard Test
PVT: Psychomotor Vigilance Task
RT: response time
SAE: serious adverse event
SDF: source data file
SNB: Spatial n-back Task
SOA: secondary organic aerosol
sp: spirometry
UREC: University of Manchester Research Ethics Committee
vs: vital signs

### Appendix A. Inclusion and exclusion criteria

The study inclusion criteria are:

- Age ≥ 50 years of age.
- Family history of dementia.
- Provision of signed, written, and dated informed consent, prior to any study specific procedures. Participants will be excluded if any of the following criteria apply:
- Neurological or psychiatric disorder.
- General Practitioner Assessment of Cognition Score of ≤ 7.
- Current smoker (including e-cigarettes) or ex-smoker of less than 6 months abstinence, and < 20 pack year history.
- Current pregnancy.
- Significantly abnormal spirometry.
- History of cardiac disease.
- History of chronic respiratory or airways disease.
- History of inflammatory diseases (e.g., rheumatoid arthritis, chronic periodontitis, inflammatory bowel disease)
- History of neurological or psychiatric disorders.
- History of organ transplant.
- Consumption of > 3 units of alcohol in the 24-hours preceding testing visit.
- Any vaccination in the week prior to the testing visit.
- Cold or flu in the 48-hours prior to the testing visit.
- Visual impairment not correctable with glasses / contact lenses.
- Significant claustrophobia preventing the participant from wearing a mask over the nose and mouth for an hour.
- Concomitant medication use of neurologically active drugs such as opiates / benzodiazepines / anti-epileptics etc.
- Substance misuse.

## References

1. World Health Organization. Ambient Air Pollution: A global assessment of exposure and burden of disease. https://www.who.int/publications/i/item/9789241511353, 2016. Accessed 10 Jan 2022.

2. Landrigan, P.J.; Fuller, R.; Acosta, N.J.R.; Adeyi, O.; Arnold, R.; Basu, N.n.; Baldé, A.B.; Bertollini, R.; Bose-O’Reilly, S.; Boufford, J.I.; et al. The Lancet Commission on pollution and health. Lancet 2018, 391, 462–512.

3. Pope, 3rd, C.A.; Dockery, D.W. Health effects of fine particulate air pollution: lines that connect. J. Air Waste Manag. Assoc. 2006, 56, 709–742.

4. Harry, G.J.; Kraft, A.D. Neuroinflammation and microglia: considerations and approaches for neurotoxicity assessment. Expert Opin. Drug Metab. Toxicol. 2008, 4, 1265–1277.

5. Costa, L.G.; Cole, T.B.; Coburn, J.; Chang, Y.C.; Dao, K.; Roqué, P.J. Neurotoxicity of traffic-related air pollution. Neurotoxicology 2017, 59, 133–139.

6. de Prado Bert, P.; Mercader, E.M.H.; Pujol, J.; Sunyer, J.; Mortamais, M. The effects of air pollution on the brain: A review of studies interfacing environmental epidemiology and neuroimaging. Curr. Environ. Health Rep. 2018, 5, 351–364.

7. Delgado-Saborit, J.M.; Guercio, V.; Gowers, A.M.; Shaddick, G.; Fox, N.C.; Love, S. A critical review of the epidemiological evidence of effects of air pollution on dementia, cognitive function and cognitive decline in adult population. Sci. Total Environ. 2021, 757, 143734.

8. Herting, M.M.; Younan, D.; Campbell, C.E.; Chen, J.C. Outdoor air pollution and brain structure and function from across childhood to young adulthood: A methodological review of brain MRI studies. Front. Public Health 2019, 7, 332.

9. Castellani, B.; Bartington, S.; Wistow, J.; Heckels, N.; Ellison, A.; Van Tongeren, M.; Arnold, S.R.; Barbrook-Johnson, P.; Bicket, M.; Pope, F.D.; et al. Mitigating the impact of air pollution on dementia and brain health: Setting the policy agenda. Environ. Res. 2022, 215, 114362.

10. Peters, R.; Ee, N.; Peters, J.; Booth, A.; Mudway, I.; Anstey, K.J. Air pollution and dementia: A systematic review. J. Alzheimers. Dis. 2019, 70, S145–S163.

11. Kasdagli, M.I.; Katsouyanni, K.; Dimakopoulou, K.; Samoli, E. Air pollution and Parkinson’s disease: A systematic review and meta-analysis up to 2018. Int. J. Hyg. Environ. Health 2019, 222, 402–409.

12. Noorimotlagh, Z.; Azizi, M.; Pan, H.F.; Mami, S.; Mirzaee, S.A. Association between air pollution and Multiple Sclerosis: A systematic review. Environ. Res. 2021, 196, 110386.

13. Delgado-Saborit.; Beevers.; Dajnak.; Church.; Gulliver.; Hoek.; Nieuwenhuijsen.; Kelly.; Sunyer.; Guxens. Air pollution exposure and cognitive and academic performance in children. Environ. Epidemiol. 2019, 3, 95.

14. Rivas, I.; Basagaña, X.; Cirach, M.; López-Vicente, M.; Suades-González, E.; Garcia-Esteban, R.; Álvarez-Pedrerol, M.; Dadvand, P.; Sunyer, J. Association between early life exposure to air pollution and working memory and attention. Environ. Health Perspect. 2019, 127, 57002.

15. Basagaña, X.; Esnaola, M.; Rivas, I.; Amato, F.; Alvarez-Pedrerol, M.; Forns, J.; López-Vicente, M.; Pujol, J.; Nieuwenhuijsen, M.; Querol, X.; et al. Neurodevelopmental deceleration by urban fine particles from different emission sources: A longitudinal observational study. Environ. Health Perspect. 2016, 124, 1630–1636.

16. Forns, J.; Dadvand, P.; Esnaola, M.; Alvarez-Pedrerol, M.; López-Vicente, M.; Garcia-Esteban, R.; Cirach, M.; Basagaña, X.; Guxens, M.; Sunyer, J. Longitudinal association between air pollution exposure at school and cognitive development in school children over a period of 3.5 years. Environ. Res. 2017, 159, 416–421.

17. Wang, S.; Zhang, J.; Zeng, X.; Zeng, Y.; Wang, S.; Chen, S. Association of traffic-related air pollution with children’s neurobehavioral functions in Quanzhou, China. Environ. Health Perspect. 2009, 117, 1612–1618.

18. Calderón-Garcidueñas, L.; Engle, R.; Mora-Tiscareño, A.; Styner, M.; Gómez-Garza, G.; Zhu, H.; Jewells, V.; Torres-Jardón, R.; Romero, L.; Monroy-Acosta, M.E.; et al. Exposure to severe urban air pollution influences cognitive outcomes, brain volume and systemic inflammation in clinically healthy children. Brain Cogn. 2011, 77, 345–355.

19. Sunyer, J.; Esnaola, M.; Alvarez-Pedrerol, M.; Forns, J.; Rivas, I.; López-Vicente, M.; Suades-González, E.; Foraster, M.; Garcia-Esteban, R.; Basagaña, X.; et al. Association between traffic-related air pollution in schools and cognitive development in primary school children: a prospective cohort study. PLoS Med. 2015, 12, e1001792.

20. Saenen, N.D.; Provost, E.B.; Viaene, M.K.; Vanpoucke, C.; Lefebvre, W.; Vrijens, K.; Roels, H.A.; Nawrot, T.S. Recent versus chronic exposure to particulate matter air pollution in association with neurobehavioral performance in a panel study of primary schoolchildren. Environ. Int. 2016, 95, 112–119.

21. Bos, I.; De Boever, P.; Vanparijs, J.; Pattyn, N.; Panis, L.I.; Meeusen, R. Subclinical effects of aerobic training in urban environment. Med. Sci. Sports Exerc. 2013, 45, 439–447.

22. Gui, Z.; Cai, L.; Zhang, J.; Zeng, X.; Lai, L.; Lv, Y.; Huang, C.; Chen, Y. Exposure to ambient air pollution and executive function among Chinese primary schoolchildren. Int. J. Hyg. Environ. Health 2020, 229, 113583.

23. Shehab, M.A.; Pope, F.D. Effects of short-term exposure to particulate matter air pollution on cognitive performance. Sci. Rep. 2019, 9, 8237.

24. Shou, Y.; Huang, Y.; Zhu, X.; Liu, C.; Hu, Y.; Wang, H. A review of the possible associations between ambient PM2.5 exposures and the development of Alzheimer’s disease. Ecotoxicol. Environ. Saf. 2019, 174, 344–352.

25. Adivi, A.; Lucero, J.; Simpson, N.; McDonald, J.D.; Lund, A.K. Exposure to traffic-generated air pollution promotes alterations in the integrity of the brain microvasculature and inflammation in female ApoE-/-mice. Toxicol. Lett. 2021, 339, 39–50.

26. Maher, B.A. Airborne magnetite- and iron-rich pollution nanoparticles: Potential neurotoxicants and environmental risk factors for neurodegenerative disease, including Alzheimer’s disease. J. Alzheimers. Dis. 2019, 71, 361–375.

27. Calderón-Garcidueñas, L.; Reynoso-Robles, R.; González-Maciel, A. Combustion and friction-derived nanoparticles and industrial-sourced nanoparticles: The culprit of Alzheimer and Parkinson’s diseases. Environ. Res. 2019, 176, 108574.

28. Seaton, A.; Tran, L.; Chen, R.; Maynard, R.L.; Whalley, L.J. Pollution, particles, and dementia: A hypothetical causative pathway. Int. J. Environ. Res. Public Health 2020, 17, 862.

29. Shou, Y.; Zhu, X.; Zhu, D.; Yin, H.; Shi, Y.; Chen, M.; Lu, L.; Qian, Q.; Zhao, D.; Hu, Y.; et al. Ambient PM2.5 chronic exposure leads to cognitive decline in mice: From pulmonary to neuronal inflammation. Toxicol. Lett. 2020, 331, 208–217.

30. Bakolis, I.; Hammoud, R.; Stewart, R.; Beevers, S.; Dajnak, D.; MacCrimmon, S.; Broadbent, M.; Pritchard, M.; Shiode, N.; Fecht, D.; et al. Mental health consequences of urban air pollution: prospective population-based longitudinal survey. Soc. Psychiatry Psychiatr. Epidemiol. 2021, 56, 1587–1599.

31. Royal College of Physicians. Every Breath We Take: The Lifelong Impact of Air Pollution. Report of a Working Party. Technical report, London: RCP, 2016. Accessed 15 Jan 2023.

32. Holgate, S.T.; Sandström, T.; Frew, A.J.; Stenfors, N.; Nordenhall, C.; Salvi, S.; Blomberg, A.; Helleday, R.; Söderberg, M. Health effects of acute exposure to air pollution. Part I: Healthy and asthmatic subjects exposed to diesel exhaust, 2003. PMID: 14738208.

33. Langrish, J.P.; Watts, S.J.; Hunter, A.J.; Shah, A.S.V.; Bosson, J.A.; Unosson, J.; Barath, S.; Lundbäck, M.; Cassee, F.R.; Donaldson, K.; et al. Controlled exposures to air pollutants and risk of cardiac arrhythmia. Environ. Health Perspect. 2014, 122, 747–753.

34. Cliff, R.; Curran, J.; Hirota, J.A.; Brauer, M.; Feldman, H.; Carlsten, C. Effect of diesel exhaust inhalation on blood markers of inflammation and neurotoxicity: a controlled, blinded crossover study. Inhal. Toxicol. 2016, 28, 145–153.

35. Crüts, B.; van Etten, L.; Törnqvist, H.; Blomberg, A.; Sandström, T.; Mills, N.L.; Borm, P.J. Exposure to diesel exhaust induces changes in EEG in human volunteers. Part. Fibre Toxicol. 2008, 5, 4.

36. Fuller, G.W.; Tremper, A.H.; Baker, T.D.; Yttri, K.E.; Butterfield, D. Contribution of wood burning to PM10 in London. Atmos. Environ. (1994) 2014, 87, 87–94.

37. Williams, M.L.; Lott, M.C.; Kitwiroon, N.; Dajnak, D.; Walton, H.; Holland, M.; Pye, S.; Fecht, D.; Toledano, M.B.; Beevers, S.D. The Lancet Countdown on health benefits from the UK Climate Change Act: a modelling study for Great Britain. Lancet Planet. Health 2018, 2, e202–e213.

38. Reyes-Villegas, E.; Bannan, T.; Le Breton, M.; Mehra, A.; Priestley, M.; Percival, C.; Coe, H.; Allan, J.D. Online chemical characterization of food-cooking organic aerosols: Implications for source apportionment. Environ. Sci. Technol. 2018, 52, 5308–5318.

39. Naseri, M.; Jouzizadeh, M.; Tabesh, M.; Malekipirbazari, M.; Gabdrashova, R.; Nurzhan, S.; Farrokhi, H.; Khanbabaie, R.; Mehri-Dehnavi, H.; Bekezhankyzy, Z.; et al. The impact of frying aerosol on human brain activity. Neurotoxicology 2019, 74, 149–161.

40. Centre for Atmospheric Science - The University of Manchester. The Manchester Aerosol Chamber. http://www.cas.manchester.ac.uk/restools/aerosolchambe, n.d. Accessed 18 Jan 2023.

41. Shao, Y.; Wang, Y.; Du, M.; Voliotis, A.; Alfarra, M.R.; O’Meara, S.P.; Turner, S.F.; McFiggans, G. Characterisation of the Manchester Aerosol Chamber facility. Atmos. Meas. Tech. 2022, 15, 539–559.

42. Hamilton, J.F.; Rami Alfarra, M.; Wyche, K.P.; Ward, M.W.; Lewis, A.C.; McFiggans, G.B.; Good, N.; Monks, P.S.; Carr, T.; White, I.R.; et al. Investigating the use of secondary organic aerosol as seed particles in simulation chamber experiments. Atmos. Chem. Phys. 2011, 11, 5917–5929.

43. Alfarra, M.R.; Hamilton, J.F.; Wyche, K.P.; Good, N.; Ward, M.W.; Carr, T.; Barley, M.H.; Monks, P.S.; Jenkin, M.E.; Lewis, A.C.; et al. The effect of photochemical ageing and initial precursor concentration on the composition and hygroscopic properties of β-caryophyllene secondary organic aerosol. Atmos. Chem. Phys. 2012, 12, 6417–6436.

44. Jenkin, M.E.; Wyche, K.P.; Evans, C.J.; Carr, T.; Monks, P.S.; Alfarra, M.R.; Barley, M.H.; McFiggans, G.B.; Young, J.C.; Rickard, A.R. Development and chamber evaluation of the MCM v3.2 degradation scheme for β-caryophyllene. Atmos. Chem. Phys. 2012, 12, 5275–5308.

45. Alfarra, M.R.; Good, N.; Wyche, K.P.; Hamilton, J.F.; Monks, P.S.; Lewis, A.C.; McFiggans, G. Water uptake is independent of the inferred composition of secondary aerosols derived from multiple biogenic VOCs. Atmos. Chem. Phys. 2013, 13, 11769–11789.

46. Wyche, K.P.; Ryan, A.C.; Hewitt, C.N.; Alfarra, M.R.; McFiggans, G.; Carr, T.; Monks, P.S.; Smallbone, K.L.; Capes, G.; Hamilton, J.F.; et al. Emissions of biogenic volatile organic compounds and subsequent photochemical production of secondary organic aerosol in mesocosm studies of temperate and tropical plant species. Atmos. Chem. Phys. 2014, 14, 12781–12801.

47. Wyche, K.P.; Monks, P.S.; Smallbone, K.L.; Hamilton, J.F.; Alfarra, M.R.; Rickard, A.R.; McFiggans, G.B.; Jenkin, M.E.; Bloss, W.J.; Ryan, A.C.; et al. Mapping gas-phase organic reactivity and concomitant secondary organic aerosol formation: chemometric dimension reduction techniques for the deconvolution of complex atmospheric data sets. Atmos. Chem. Phys. 2015, 15, 8077–8100.

48. Pereira, K.L.; Dunmore, R.; Whitehead, J.; Alfarra, M.R.; Allan, J.D.; Alam, M.S.; Harrison, R.M.; McFiggans, G.; Hamilton, J.F. Technical note: Use of an atmospheric simulation chamber to investigate the effect of different engine conditions on unregulated VOC-IVOC diesel exhaust emissions. Atmos. Chem. Phys. 2018, 18, 11073–11096.

49. Ting, Y.; Mitchell, E.J.S.; Allan, J.D.; Liu, D.; Spracklen, D.V.; Williams, A.; Jones, J.M.; Lea-Langton, A.R.; McFiggans, G.; Coe, H. Mixing state of carbonaceous aerosols of primary emissions from “improved” African cookstoves. Environ. Sci. Technol. 2018, 52, 10134–10143.

50. Mitchell, E.J.S.; Ting, Y.; Allan, J.; Lea-Langton, A.R.; Spracklen, D.V.; McFiggans, G.; Coe, H.; Routledge, M.N.; Williams, A.; Jones, J.M. Pollutant emissions from improved cookstoves of the type used in sub-Saharan Africa. Combust. Sci. Technol. 2020, 192, 1582–1602.

51. Stewart, G.J.; Nelson, B.S.; Acton, W.J.F.; Vaughan, A.R.; Farren, N.J.; Hopkins, J.R.; Ward, M.W.; Swift, S.J.; Arya, R.; Mondal, A.; et al. Emissions of intermediate-volatility and semi-volatile organic compounds from domestic fuels used in Delhi, India.

52. Stewart, G.J.; Acton, W.J.F.; Nelson, B.S.; Vaughan, A.R.; Hopkins, J.R.; Arya, R.; Mondal, A.; Jangirh, R.; Ahlawat, S.; Yadav, L.; et al. Emissions of non-methane volatile organic compounds from combustion of domestic fuels in Delhi, India.

53. Graham, B.L.; Steenbruggen, I.; Miller, M.R.; Barjaktarevic, I.Z.; Cooper, B.G.; Hall, G.L.; Hallstrand, T.S.; Kaminsky, D.A.; McCarthy, K.; McCormack, M.C.; et al. Standardization of spirometry 2019 update. An official American Thoracic Society and European Respiratory Society technical statement. Am. J. Respir. Crit. Care Med. 2019, 200, e70–e88.

54. Brodaty, H.; Pond, D.; Kemp, N.M.; Luscombe, G.; Harding, L.; Berman, K.; Huppert, F.A. The GPCOG: a new screening test for dementia designed for general practice. J. Am. Geriatr. Soc. 2002, 50, 530–534.

55. Tiffin, J.; Asher, E.J. The Purdue pegboard; norms and studies of reliability and validity. J. Appl. Psychol. 1948, 32, 234–247.

56. The MathWorks Inc. Matlab version: 9.12 (R2022a), Natick, Massachusetts: The MathWorks Inc. 2022. http://www.mathworks.com, n.d.

57. Brainard, D.H. The Psychophysics Toolbox. Spat. Vis. 1997, 10, 433–436.

58. Chan, A.W.; Tetzlaff, J.M.; Gøtzsche, P.C.; Altman, D.G.; Mann, H.; Berlin, J.A.; Dickersin, K.; Hróbjartsson, A.; Schulz, K.F.; Parulekar, W.R.; et al. SPIRIT 2013 explanation and elaboration: guidance for protocols of clinical trials. BMJ 2013, 346, e7586.

