## Supplementary File 1 - Informed Consent Form for "HIPTox – Hazard Identification Platform to Assess the Health Impacts from Indoor and Outdoor Air Pollutant Exposures, through Mechanistic Toxicology: A single-centre double-blind human exposure trial protocol"

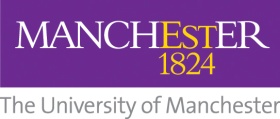

HIPTOX study:

Hazard Identification Platform to Assess the Health Impacts from Indoor and Outdoor Air Pollutant Exposures, through Mechanistic Toxicology; Human Studies

**Consent Form Version 1.5 23^rd^ February 2023**

If you are happy to participate please complete and sign the consent form below.

|  | **Activities** | Initials |
| --- | --- | --- |
| 1 | I confirm that I have read the attached information sheet (Version 1.6 dated 23^rd^ February 2023) for the above study and have had the opportunity to consider the information and ask questions and had these answered satisfactorily. |  |
| 2 | I understand that my participation in the study is voluntary and that I am free to withdraw at any time without giving a reason and without detriment to myself. I understand that it will not be possible to remove my data from the project once it has been anonymised and forms part of the data set.  I agree to take part on this basis. |  |
| 3 | I agree to my GP being informed of my participation in this study. |  |
| 4 | I agree to have **blood samples and nasal washings** taken for the research purpose as explained to me. |  |
| 5 | I agree that any data collected may be included in anonymous form in **publications/conference presentations.** |  |
| 6 | I agree that any anonymised data collected may be made available to other researchers |  |
| 7 | I understand that data collected during the study may be looked at by individuals from The University of Manchester or regulatory authorities, where it is relevant to my taking part in this research. I give permission for these individuals to have access to my data. |  |
| 8 | I understand that there may be instances where during the course of the research information is revealed which means the researchers will be obliged to break confidentiality and this has been explained in more detail in the information sheet. |  |
| 9 | I agree to take part in this study. |  |

**The following activities are optional, you may participate in the research without agreeing to the following:**

| 10 | I understand that the sponsors of this study may make my **blood sample** available to other researchers **for future research**. I understand that this will involve **genetic research**. I give permission for these individuals to have access to my **sample, (but not any personal identifying information about me,) I offer my blood sample as a gift.** |
| --- | --- |
| 11 | I agree that the researchers may retain my contact details to provide me with a summary of the findings for this study. |

**Data Protection**

**The personal information we collect and use to conduct this research will be processed in accordance with data protection law as explained in the Participant Information Sheet and the** [**Privacy Notice for Research Participants**](http://documents.manchester.ac.uk/display.aspx?DocID=37095)

________________________	________________________

Name of Participant	Signature	Date

________________________	________________________

Name of the person taking consent	Signature	Date

[1 copy for the participant, 1 copy for the research team (original), 1 copy for the medical notes]
