## Supplementary File 2 - GPCog for "HIPTox – Hazard Identification Platform to Assess the Health Impacts from Indoor and Outdoor Air Pollutant Exposures, through Mechanistic Toxicology: A single-centre double-blind human exposure trial protocol"

Patient name: ___________________________

Testing date: ___________________________

GPCOG

### STEP 1 – PATIENT EXAMINATION

Unless specified, each question should only be asked once.

#### Name and address for subsequent recall test

*I am going to give you a name and address. After I have said it, I want you to repeat it. Remember this name and address because I am going to ask you to tell it to me again in a few minutes: John Brown, 42 West Street, Kensington.* (Allow a maximum of 4 attempts.)

| | Correct | Incorrect |
|---|---|---|
| **Time orientation** | | |
| 1. *What is the date?* (exact only) | ☐ | ☐ |
| **Clock drawing** (use blank page) | | |
| 2. *Please mark in all the numbers to indicate the hours of a clock.* (correct spacing required) | ☐ | ☐ |
| 3. *Please mark in hands to show 10 minutes past eleven o'clock.* (11.10) | ☐ | ☐ |
| **Information** | | |
| 4. *Can you tell me something that happened in the news recently?* (Recently = in the last week. If a general answer is given, e.g. "war", "lot of rain", ask for details. Only specific answer scores.) | ☐ | ☐ |
| **Recall** | | |
| 5. *What was the name and address I asked you to remember?* | | |
| John | ☐ | ☐ |
| Brown | ☐ | ☐ |
| 42 | ☐ | ☐ |
| West (St) | ☐ | ☐ |
| Kensington | ☐ | ☐ |
| Add the number of items answered correctly: **Total score:** | ☐ | **out of 9** |

| | |
|---|---|
| **9** | **No significant cognitive impairment**<br>Further testing is not necessary |
| **5 – 8** | **More information required**<br>Proceed with informant interview in step 2 on next page |
| **0 – 4** | **Cognitive impairment is indicated**<br>Conduct standard investigations |

© University of New South Wales as represented by the Dementia Collaborative Research Centre – Assessment and Better Care; Brodaty et al, *JAGS* 2002; 50:530-534
