## Supplementary File 3 - Previous Chamber Study Safety for "HIPTox – Hazard Identification Platform to Assess the Health Impacts from Indoor and Outdoor Air Pollutant Exposures, through Mechanistic Toxicology: A single-centre double-blind human exposure trial protocol"

**Supplementary File 3 – Summary of human chamber studies (limited to diesel and woodsmoke exposures)**

| **Studies** | **Exposures** | **Duration** | **Number of subjects** | **Procedures** | **Adverse responses** |
| --- | --- | --- | --- | --- | --- |
| [Salvi et al., 1999](https://doi.org/10.1164/ajrccm.159.3.9709083) | Diesel - 300 μg m^‑3^ vs. filtered air. | 1 hour – with alternate 15 mins of exercise and rest | Healthy Controls | Spirometry, bronchoscopy – lavage and biopsy | None |
| [Nordenhäll et al., 2001](https://doi.org/10.1183/09031936.01.17509090) | Diesel - 300 μg m^‑3^ vs. filtered air. | 1 hour – with alternate 15 mins of exercise and rest | Asthmatics: 14 | Induced sputum + airway hyperresponsiveness | None |
| [Holgate et al., 2003](https://pubmed.ncbi.nlm.nih.gov/14738208/) | Diesel - 100 μg m^‑3^ vs. filtered air. | 2 hours – with alternate 15 mins of exercise and rest | Healthy Controls: 25  Asthmatics: 15 | Spirometry, bronchoscopy – lavage and biopsy | None |
| [Mills et al., 2005](https://doi.org/10.1161/CIRCULATIONAHA.105.588962) | Diesel - 300 μg m^‑3^ vs. filtered air. | 1 hour – with alternate 15 mins of exercise and rest | Healthy Controls: 30 | plethysmography | None |
| [Mills et al., 2007](https://doi.org/10.1056/NEJMoa066314) | Diesel - 300 μg m^‑3^ vs. filtered air. | 1 hour – with alternate 15 mins of exercise and rest | Patients with prior myocardial infarction: 20 | Electrocardiography, plethysmography | None |
| [Sehlstedt et al., 2010](https://doi.org/10.1186/1743-8977-7-21) | Woodsmoke 200 μg m^‑3^ vs. filtered air. | 3 hours – with alternate 15 mins of exercise and rest | Healthy Controls: 19 | Spirometry, bronchoscopy – lavage and biopsy | None |
| [Behndig et al., 2011](https://doi.org/10.1136/thx.2010.140053) | Diesel - 100 μg m^‑3^ vs. filtered air. | 2 hours – with alternate 15 mins of exercise and rest | Healthy Controls: 23  Asthmatics: 32 | Spirometry, bronchoscopy – lavage and biopsy | None |
| [Mills & Finlayson et al., 2011](https://doi.org/10.1136/hrt.2010.199042) | Diesel - 300 μg m^‑3^ vs. filtered air. | 1 hour – with alternate 15 mins of exercise and rest | Healthy Controls: 32  Patients with prior myocardial infarction: 20 | Electrocardiography | None |
| [Mills & Miller et al., 2011](https://doi.org/10.1093/eurheartj/ehr195) | Diesel - 300 μg m^‑3^ Carbon nanoparticles - 70 μg m^‑3^  Filtered diesel exhaust – 6 μg m^‑3^  filtered air | 2 hours – with alternate 15 mins of exercise and rest | Healthy Controls: 16 | Plethysmography | None |
| [Unosson et al., 2013](https://doi.org/10.1186/1743-8977-10-20) | Wood smoke – 300 μg m^‑3^ vs. filtered air. | 3 hours – with alternate 15 mins of exercise and rest | Healthy Controls: 16 | Electrocardiography, plethysmography | None |
| [Langrish et al., 2013](https://doi.org/10.1161/JAHA.112.004309) | Diesel - 300 μg m^‑3^ vs. filtered air. | 1 hour – with alternate 15 mins of exercise and rest | Healthy Controls: 16 (Study 1) | Plethysmography | None |
| [Muala et al., 2015](https://doi.org/10.1186/s12989-015-0111-7) | Woodsmoke - 300 μg m^‑3^ vs. filtered air. | 3 hours – with alternate 15 mins of exercise and rest | Healthy controls: 14 | Spirometry, bronchoscopy – lavage and biopsy | None |
| [Hunter et al., 2014](https://doi.org/10.1186/s12989-014-0062-4) | Woodsmoke - 1000 μg m^‑3^ vs. filtered air | 1 hour – with alternate 15 mins of exercise and rest | Healthy Controls: 16 | Plethysmography | None |
| [Carlsten et al., 2016](https://doi.org/10.1136/thoraxjnl-2015-207399) | Diesel – 300 μg m^‑3^ vs. filtered air.  Followed by segmental allergen challenge. | 1 hour – with alternate 15 mins of exercise and rest | Atopic adults | Spirometry, bronchoscopy – lavage and biopsy | None |
| [Rankin et al., 2021](https://doi.org/10.1161/JAHA.120.018448) | Diesel - 300 μg m^‑3^ vs. filtered air. | 1 hour – with alternate 15 mins of exercise and rest | Healthy Controls: 16 | Microneurography | None |
| [Unosson et al., 2021](https://doi.org/10.1186/s12989-021-00412-3) | Study 1: Diesel - 300 μg m^‑3^ vs.  Biodiesel - 30% RME - 300 μg m^‑3^  Study 2:  Diesel – 300 μg m^‑3^ vs.  Biodiesel - 100% RME - 165 μg m^‑3^ | 1 hour – with alternate 15 mins of exercise and rest | Healthy Controls  Study 1: 16  Study 2: 19 | Plethysmography | None |

**References**

Behndig, A. F., Larsson, N., Brown, J. L., ... (2011). Proinflammatory doses of diesel exhaust in healthy subjects fail to elicit equivalent or augmented airway inflammation in subjects with asthma. Thorax, 66(1), 12-19.

Carlsten, C., Blomberg, A., Pui, M., ... (2016). Diesel exhaust augments allergen-induced lower airway inflammation in allergic individuals: a controlled human exposure study. Thorax, 71(1), 35-44. <https://doi.org/10.1136/thoraxjnl-2015-207399> PMID: 265

Holgate, S. T., Sandström, T., Frew, A. J., Stenfors, N., Nordenhall, C., Salvi, S., ... & Söderberg, M. (2003). Health effects of acute exposure to air pollution. Part I: Healthy and asthmatic subjects exposed to diesel exhaust. PMID: 14738208.

Hunter, A. L., Unosson, J., Bosson, J. A., ... (2014). Effect of wood smoke exposure on vascular function and thrombus formation in healthy fire fighters. Particle and Fibre Toxicology, 11(1), 62. <https://doi.org/10.1186/s12989-014-0062-4>

Langrish, J. P., Unosson, J., Bosson, J., ... (2013). Altered nitric oxide bioavailability contributes to diesel exhaust inhalation-induced cardiovascular dysfunction in man. *Journal of the American Heart Association, 2*(1), e004309. <https://doi.org/10.1161/JAHA.112.004309> PMID: 23525434; PMCID: PMC3603248.

Mills, N. L., Finlayson, A. E., Gonzalez, M. C., Törnqvist, H., Barath, S., Vink, E., ... & Newby, D. E. (2011). Diesel exhaust inhalation does not affect heart rhythm or heart rate variability. *Heart, 97*(7), 544-550. DOI: [10.1136/hrt.2010.199042](https://doi.org/10.1136/hrt.2010.199042) PMID: 20962342.

Mills, N. L., Miller, M. R., Lucking, A. J., Beveridge, J., Flint, L., Boere, A. J., ... & Newby, D. E. (2011). Combustion-derived nanoparticulate induces the adverse vascular effects of diesel exhaust inhalation. European Heart Journal, 32(21), 2660-2671. <https://doi.org/10.1093/eurheartj/ehr195> PMID: 21753226; PMCID: PMC3205591.

Mills, N. L., Törnqvist, H., Gonzalez, M. C., Vink, E., Robinson, S. D., Söderberg, S., ... & Newby, D. E. (2007). Ischemic and thrombotic effects of dilute diesel-exhaust inhalation in men with coronary heart disease. New England Journal of Medicine, 357(11), 1075-1082. <https://doi.org/10.1056/NEJMoa066314> PMID: 17855668.

Mills, N. L., Törnqvist, H., Robinson, S. D., Gonzalez, M., Darnley, K., MacNee, W., ... & Newby, D. E. (2005). Diesel exhaust inhalation causes vascular dysfunction and impaired endogenous fibrinolysis. Circulation, 112(25), 3930-3936. <https://doi.org/10.1161/CIRCULATIONAHA.105.588962> PMID: 16365212.

Muala, A., Rankin, G., Sehlstedt, M., ... (2015). Acute exposure to wood smoke from incomplete combustion - indications of cytotoxicity. Particle and Fibre Toxicology, 12, 33. <https://doi.org/10.1186/s12989-015-0111-7>

Nordenhäll, C., Pourazar, J., Ledin, M-C., Levin, J-O., Sandström, T., & Ädelroth, E. (2001). European Respiratory Journal, 17(5), 909-915. <https://doi.org/10.1183/09031936.01.17509090>

Rankin, G. D., ... (2021). Acute exposure to diesel exhaust increases muscle sympathetic nerve activity in humans. Journal of the American Heart Association, 10(10). <https://doi.org/10.1161/jaha.120.018448>

Salvi, S., Blomberg, A., Rudell, B., Kelly, F., Sandström, T., Holgate, S. T., ... & Frew, A. (1999). Acute inflammatory responses in the airways and peripheral blood after short-term exposure to diesel exhaust in healthy human volunteers. American Journal of Respiratory and Critical Care Medicine, 159(3), 702-709. <https://doi.org/10.1164/ajrccm.159.3.9709083> PMID: 10051240.

Sehlstedt, M., Dove, R., Boman, C., Pagels, J., Swietlicki, E., Löndahl, J., ... & Blomberg, A. (2010). Antioxidant airway responses following experimental exposure to wood smoke in man. Particle and Fibre Toxicology, 7, 21. <https://doi.org/10.1186/1743-8977-7-21> PMID: 20727160; PMCID: PMC2936868.

Unosson, J., Blomberg, A., Sandström, T., ... (2013). Exposure to wood smoke increases arterial stiffness and decreases heart rate variability in humans. Particle and Fibre Toxicology, 10, 20. <https://doi.org/10.1186/1743-8977-10-20>

Unosson, J., Kabéle, M., Boman, C., ... (2021). Acute cardiovascular effects of controlled exposure to dilute Petrodiesel and biodiesel exhaust in healthy volunteers: a crossover study. Particle and Fibre Toxicology, 18(1), 22. <https://doi.org/10.1186/s12989-021-00412-3> PMID: 34127003; PMCID: PMC8204543.
