## Supplementary File 4 - Cognitive Task Methodology for "HIPTox – Hazard Identification Platform to Assess the Health Impacts from Indoor and Outdoor Air Pollutant Exposures, through Mechanistic Toxicology: A single-centre double-blind human exposure trial protocol"

A Windows 10 computer running Matrix Laboratory (MATLAB) version R2022a (The MathWorks Inc, 2022) will be used to run the cognitive tasks. All tasks are in the format of a MATLAB script utilising the Psychophysics Toolbox version 3.0.18 ([Brainard, 1997](https://doi.org/10.1163/156856897X00357)).

**Spatial *n*-back Task**

This spatial memory task broadly tested the encoding and immediate retrieving of temporary memory (working memory). The *n*-back task is a continuous performance task used to measure working memory capacity ([Kirchner, 1958](https://doi.org/10.1037/h0043688)). In this common task, participants are presented with stimulus sequences and must decide if the currently presented stimulus matches the stimulus presented ‘*n*’ trials ago. As ‘*n*’ increases, so does the amount of information stored in working memory. It is widely acknowledged that working memory is limited, with [Miller (1956)](https://doi.org/10.1037/h0043158) suggesting the number of items that can be held in working memory is 7 ± 2. Therefore, as ‘*n*’ increases, the difficulty of the task, and proportion of errors, increases.

***Stimuli***

A 3x3 centrally presented grid cross was used. The background colour was grey (red-green-blue coordinate, RGB [128, 128, 128]) and explanatory text and fixation crosses appeared in white, RGB [255, 255, 255].

***Procedure***

See Figure SNB (A) for the sequence of displays in each trial. For each trial, a blank 3x3 grid is presented for 600 ms followed by a one white square presented in one of the nine available grid locations for 1,000 ms. Participants are then shown the blank grid once again. Participants are tasked with remembering the sequence of grid locations and respond on *each* trial as to whether the square is in the same location as ‘*n*’ trials previously, or different. An example of this for ‘*n*’ as 1 and 2 respectively can be seen in Figure SNB (B). Participants respond using the “m” and “z” keyboard keys for ‘same’ and ‘different’ respectively. There are three blocks of 45 trials, each containing 8 matches i.e., ‘same’ locations. The *n* value increased each block of trials, i.e., block 1, n = 1; block 3, *n* = 3.

Working memory ability is indexed by d′, the standardized difference between the means of the Signal Present (same location) and Signal Absent (different location) distributions. This is calculated as Z-score Hit Rate [#Hits / (#Hits + #Misses) minus Z-score False Alarm Rate (#False Alarms / (#False Alarms + #Correct Rejections)]. A low score indicates poorer working memory ability. Due to the aforementioned information about working memory load, it is expected that as ‘*n*’ increases, performance will decrease.

Response Time (RT) will not be used as a metric of interest, as participants are given a relatively long time (10 seconds) to respond to whether the stimulus is the same or different as ‘*n*’ trials previously.

**Figure SNB**

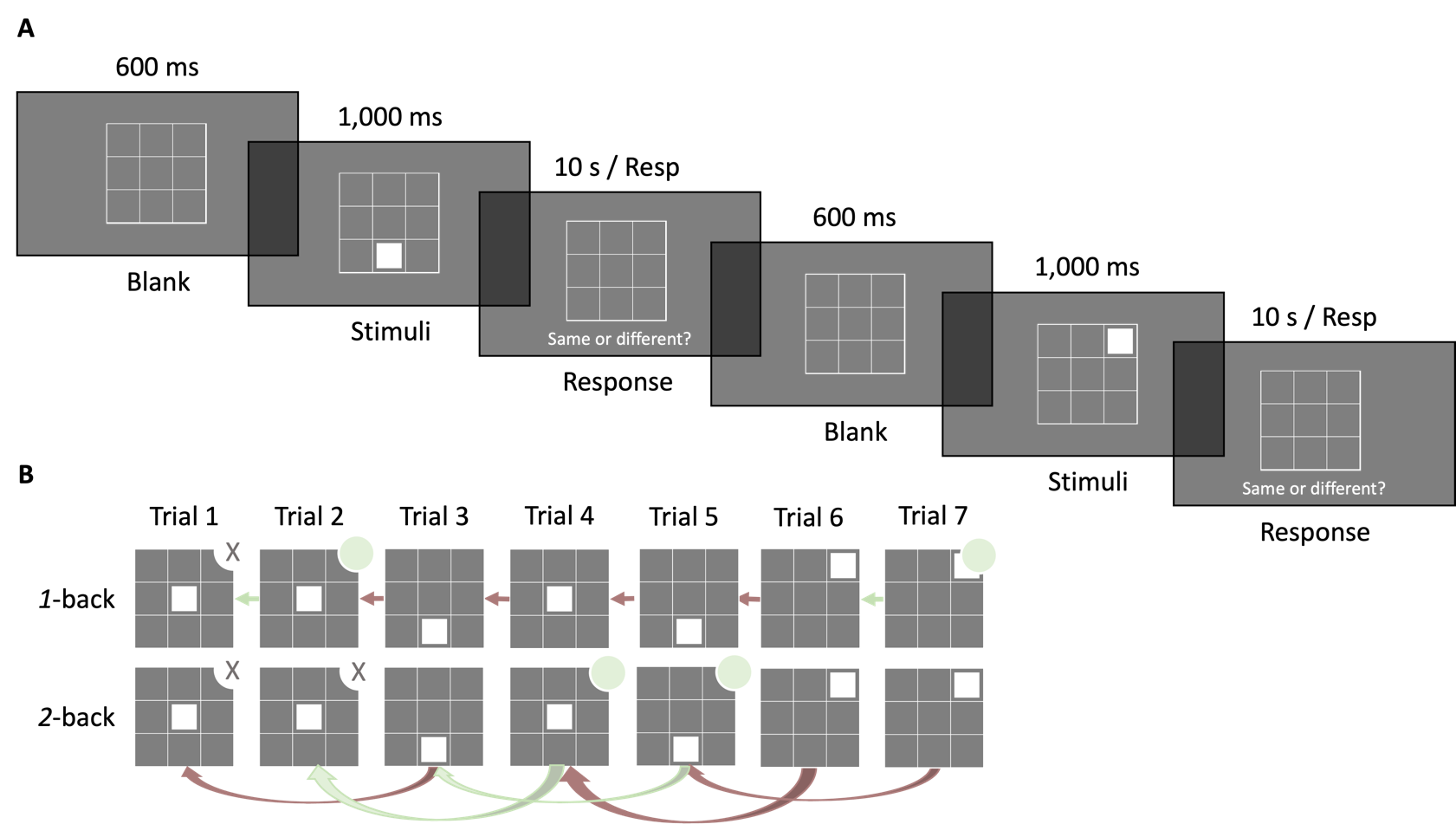
*Illustration of the Spatial n-back Task and Example of 1-back and 2-back*

*Note.* **A**: Each trial begins with a blank grid for 600 milliseconds (ms) followed by the location of the white square stimuli (1,000 ms), then a response screen with the blank square for 10 seconds or until response. **B**: Example of ‘same’ responses for a 1-back and 2-back trial with the same location presentation sequence. ‘Same’ responses indicated with pale green circle in top right corner. No response would be required for trials with ‘X’ as there are not enough previous trials for comparison.

**Face Identification Task**

This task is a selective attention task, measuring broadly the same functions as a Stroop ([Stroop, 1935](https://doi.org/10.1037/h0054651)) or Flanker ([Eriksen and Eriksen, 1974](https://doi.org/10.3758/BF03203267)) task, where participants must ignore distracting information to focus on task goals (executive function).

As high-level cognitive systems are limited in capacity, the brain uses a *proactive* control mechanism to plan strategically and enable selective engagement with expected, pertinent information as well as active avoidance of predictable but distracting information. However, a *reactive* control mechanism is also available for controlling behaviour when sudden, unpredictable events occur. For example, planning to make a coffee requires proactive control to identify and walk towards the kettle, whilst avoiding the biscuit tin (because you’re on a diet!), but reactive control might be needed to avoid colliding with a suddenly appearing colleague on the way. These two mechanisms are thought to compete for control over attention with completion of planned tasks dependent on sustained proactive control ([Botvinick et al., 1999](https://doi.org/10.1038/46035); [Braver, 2012](https://doi.org/10.1016/j.tics.2011.12.010)) and distraction by unexpected, task-irrelevant events reflecting reactive control.

***Stimuli***

A white spatial cue arrow pointing left or right and a white centrally presented fixation cross are used on a black background, RGB [0, 0, 0]. Faces are from the A set of the Karolinska Directed Emotional Faces (Lundqvist et al., 1998) utilising the frightened (fearful) & smiling (happy) emotional stimuli. Scrambled images were created by splitting face images into 13,984 squares and randomising their position. Each target/distractor image subtends 9° x 12.1°, with the centre of each presented 8.8° of visual angle laterally to the left and right of centre.

***Procedure***

See Figure FIT for the sequence of displays in each trial. Participants are instructed to respond as quickly as possible identifying the target stimulus gender presentation using the ‘l’ and ‘p’ keyboard keys with index and middle fingers of their dominant hand; key assignment to ‘male’ and ‘female’ will be counterbalanced between participants. There are 4 blocks containing 60 trials each, for a total of 240. Second fixation cross is the duration the product of a random integer chosen between 20 & 50 by frame rate (17 Hz); the target array consists of one image either side of the fixation cross, for 75 ms; and a final fixation cross presented for 1,500 ms or until participants respond. The target array comprised a central fixation, distractor image (scrambled, happy, fearful), and target image (happy, fearful), with gender and emotion expression congruency balanced across all trials, making all stimuli combinations equally likely. After the short (75 ms) presentation of the target array, participants are instructed to identify the gender of the target face as quickly and accurately as possible.

**Figure FIT**

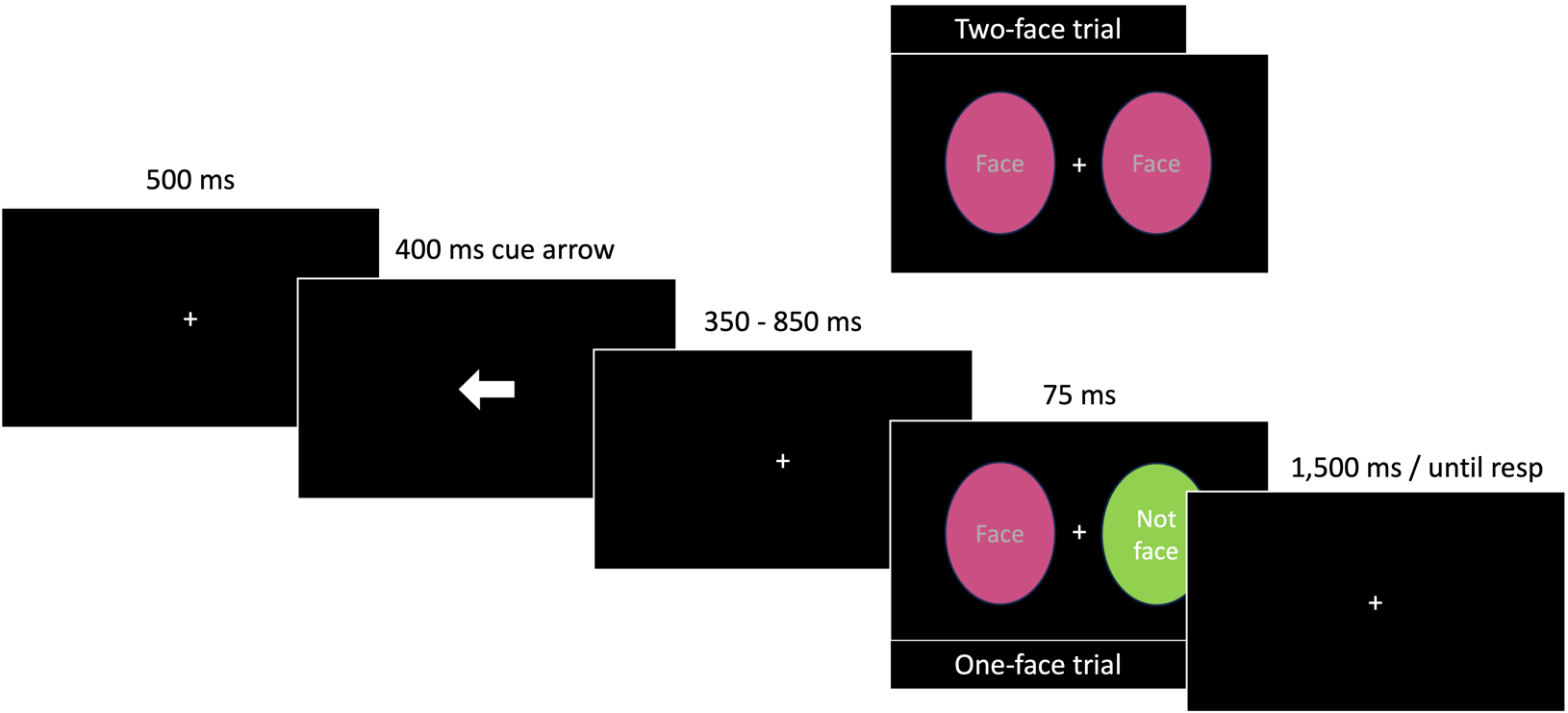
*Illustration of the Face Identification Task*

*Note.* Each trial began with a fixation cross presented for 500 milliseconds (ms), followed by a cue arrow (400 ms), a jittered fixation cross (350 – 850 ms), a quick (75 ms) presentation of the target array, and finally ended in a fixation cross awaiting response. The task was to respond as quickly as possible reporting the gender presentation of the target face, ignoring the distractor image which was either a scrambled image (one-face trial) or another face (two-face trial).

The Face Identification Task requires fast, accurate target face gender identification in the presence of another face (2-face trials) or a non-face distractor (1-face trials). Typically, in this task a face distractor slows response time indicating that it has captured reactive selective attention. Numerous studies of cognitive control show that distractor-induced slowing is exacerbated when the preceding trial has no compelling distractor, as in a 1-face trial, compared to when a distractor is present, as in a 2-face trial ([Gratton et al., 1992](https://doi.org/10.1037/0096-3445.121.4.480)). This is due to prior experience of distractor suppression on trial *n*-1 boosting proactive target processing on trial *n* whereas, when trial *n*-1 does not require suppression, proactive control is weakened leading to increased susceptibility to attention capture by an irrelevant distractor on trial *n* ([Egner and Hirsch, 2005](https://doi.org/10.1038/nn1594)). Differences in response time (RT) for 2-face trials preceded by a 2-face trial (repeat sequence) versus RT on 2-face trials preceded by a 1-face trial (change sequence) serves to inversely index proactive cognitive control, i.e., the capacity to maintain strong selection bias for the target, despite recent sensory events. It is therefore expected that in normal functioning RTs would be slower for change versus repeat sequences.

The key metric of interest in this task is cognitive control. Cognitive control will be indexed by ΔRT (RT for repeat sequences minus RT for change sequences). For clarity, “accuracy” of responses to the task goal (identifying the gender presentation of the faces) is irrelevant and was chosen to ensure the task contained a goal, but that the goal itself will not interfere with outcome metrics to infer cognitive control.

**Purdue Pegboard Test**

This is a standardised task to measure perceptual-motor function. The full task procedure and normative data is described in the manual ([Lafayette Instrument, 2023](https://lafayetteinstrument.com/downloads/manuals/MAN-32020A-pdf-rev3.pdf)).

**Expression Recognition Task**

This Go/No-go task utilising either happy or fearful expressions as the target stimuli tests the decision-making ability between positive-affective and negative-affective expressions (executive function / socio-emotional cognition).

***Stimuli***

A black centrally presented fixation cross is used. Expressive faces used in this task are from the RADIATE database ([Conley et al., 2018](https://doi.org/10.1016/j.psychres.2018.04.066)) Each face image is presented centrally. The background colour is white, RGB [255, 255, 255] and explanatory text and fixation crosses appear in black, RGB [0, 0, 0].

***Procedure***

See Figure ERT for the sequence of displays in each trial. For each block participants are instructed to respond to faces displaying the target expression (happy or fearful) and inhibit response to the other expression (fearful or happy, respectively). Participants respond as quickly as possible by pressing the keyboard spacebar using their dominant hand. Target expression is alternated across successive blocks, starting with “happy”. For each trial, a fixation cross is presented for 700 ms; followed by an expressive face image for 100 ms, or until response; followed by a blank screen for 700 ms, or until response. Four blocks of 44 trials are presented, each containing 28 target (Go) images and 16 non-target (No-go) images. Stimuli will have either open mouths (more expressive) or closed mouths (less expressive), split equally between Go and No-go trials in each block.

There are therefore four outcomes of each trial ([Harvey, 1992](https://doi.org/10.1016/0749-5978(92)90063-d)): a hit (correct “Go” response to target expression); correct rejection (correct “No-go” inhibitory response to non-target expression); miss (incorrect “No-go” to target expression); and a false alarm (incorrect “Go” response to non-target expression). See Table ERT.

**Table ERT**

*Contingency Table showing all possible Trial Outcomes*

|  |  | Stimulus Signal | |
| --- | --- | --- | --- |
|  |  | Present | Absent |
| Participant Action | Response (Go) | Hit | False Alarm |
|  | Inhibition (No-go) | Miss | Correct Rejection |

The metric d′, a measure of sensitivity to expression, is indexed by Z-score Hit Rate [#Hits / (#Hits + #Misses) minus Z-score False Alarm Rate (#False Alarms / (#False Alarms + #Correct Rejections)]. A low score indicates less sensitivity to, i.e., a greater difficulty in distinguishing, a stimulus signal.

**Figure ERT**

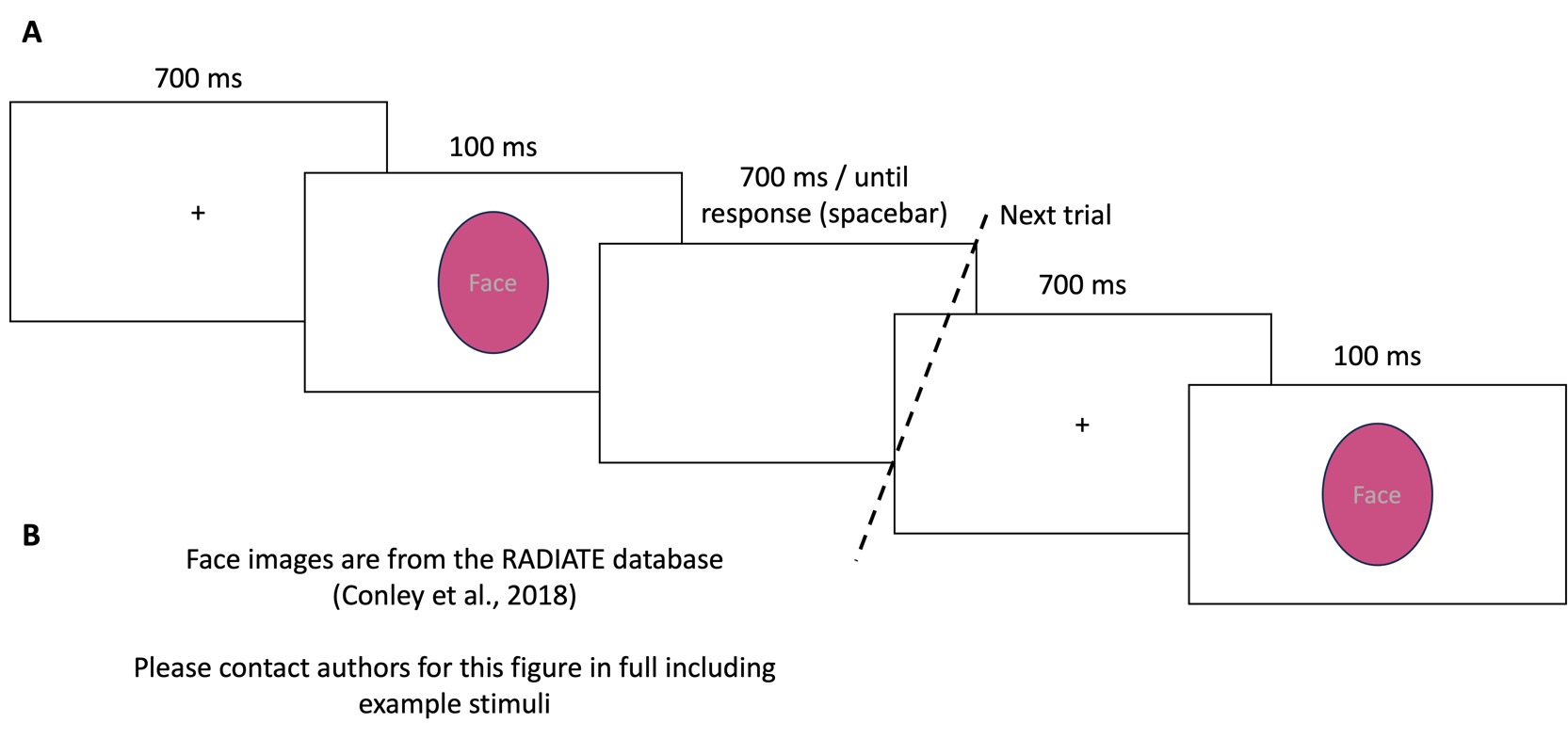
*Illustration of the Expression Recognition Task and Example stimuli*

*Note.* **A**: Each trial begins with a fixation cross presented for 700 milliseconds (ms), followed by the target array for 100 ms or until participants respond, then a blank array for 700 ms or until participants respond. The task is to respond as quickly as possible (spacebar press) if the face matches the instructed expression or inhibit response if the expression differs.

**B**: Contact authors for example positive-affective (happy) and negative-affective (fearful) stimuli.

**Psychomotor Vigilance Task**

This basic response time task tests the ability of participants to respond as quickly as possible to a change in the visual field. This task measures both simple reaction time indicative of global processing (psychomotor speed) and concentration for relatively long periods (sustained attention).

***Stimuli***

A red centrally presented circle is used RGB [255, 60, 0] against a black background RGB [0, 0, 0]. Explanatory text and fixation crosses appear in white, RGB [255, 255, 255].

***Procedure***

See Figure PVT for the sequence of displays in each trial. Participants are instructed to focus on the fixation cross positioned centrally on the screen and respond as quickly as possible when a central target (small red circle) appears. For each trial, a fixation cross is presented for between 400 and 1,800 ms (psychomotor speed trials) or between 25 and 35 seconds (sustained attention trials); followed by a target circle for 400 ms, or until response; followed by a white circle for 400 ms if spacebar is pressed during target presentation. One block of 85 trials will be presented, each containing 10 sustained attention (long fixation) trials and 75 psychomotor speed (short fixation) trials.

**Figure PVT**

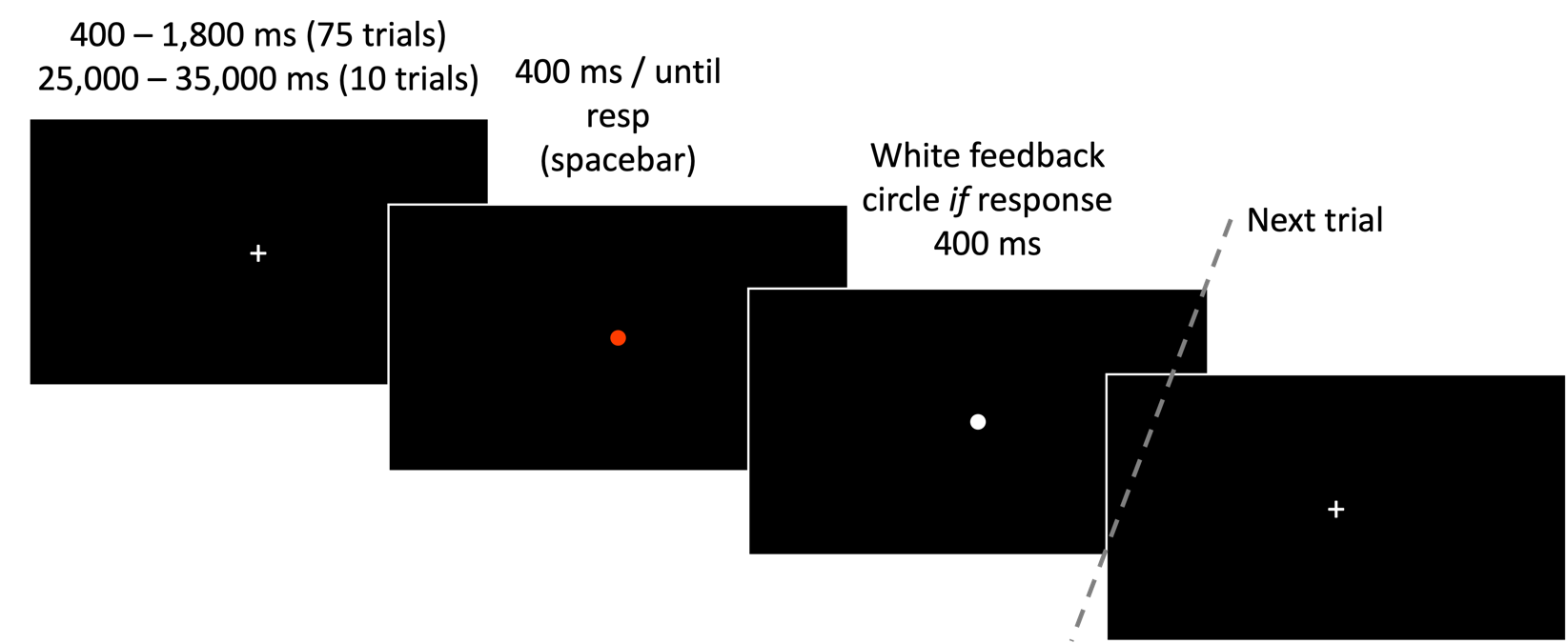
*Illustration of the Psychomotor Vigilance Task*

*Note.* Each trial begins with a fixation cross presented for between 400 and 1,800 milliseconds (psychomotor speed trials) or between 25 and 35 seconds (sustained attention trials), followed by the target dot array for 400 ms, then a feedback array if participants respond (400 ms). The task is to respond as quickly as possible (spacebar press) as soon as the target appears on the screen.

Response time (RT), regardless of if participants respond within the 400 ms, is the metric under investigation i.e., time taken between perceiving the target visual stimulus and pressing the response button. A lower RT will indicate quicker psychomotor speed and better sustained attention respectively.

It is expected that number of lapses and mean RT **will not** significantly differ between the pre-exposure and post-exposure test administrations. Indicating that exposure to low-quality air, comparative to clean, does not influence sustained attention or basic psychomotor functioning.

If significant differences are identified, this may have implications for the results of the other tasks i.e., identified RT differences may be explained simply by fatigue as opposed to more involved mechanisms linking air pollution exposure to diminished higher-order (more demanding) cognitive functioning.

**Data Analyses**

*Spatial n-back Task*: The metric d′, a measure of accuracy, will be indexed by Z-score Hit Rate [#Hits / (#Hits + #Misses) minus Z-score False Alarm Rate (#False Alarms / (#False Alarms + #Correct Rejections)]; calculated separately for each *n*-back value.

*Face Identification Task*: Trials will be excluded from statistical analyses if there was no response on the current trial or the response too fast (RT < 200 ms) on the current trial (trial *n*). Individual RTs will be trimmed if ±2 SDs from mean of change and repeat sequences respectively.

*Purdue Pegboard Test*: Gross movement performance will be indexed by Right+Left+Both score. Fine motor extremity performance will be indexed by assembly score.

*Emotional Discrimination Task*: Trials will be removed if response times (RT) are below 200 ms, indicating an anticipation error. The metric d′, a measure of expression sensitivity, will be indexed by Z-score Hit Rate [#Hits / (#Hits + #Misses) minus Z-score False Alarm Rate (#False Alarms / (#False Alarms + #Correct Rejections)]. This will be calculated separately for each emotion expression type and individual Hit RTs trimmed if ±2 SDs from mean of each.

*Psychomotor Vigilance Task*: Individual trials will be removed if response times are below 100 ms, indicating an anticipation error. Sustained attention will be indexed by RT following long fixation pauses (10 trials). Psychomotor Speed will be indexed by RT following short fixation pauses (75 trials).

**References**

Botvinick, M., Nystrom, L. E., Fissell, K., Carter, C. S., & Cohen, J. D. (1999). Conflict monitoring versus selection-for-action in anterior cingulate cortex. *Nature*, *402*, 179–181. <https://doi.org/10.1038/46035>

Brainard, D. H. (1997). The Psychophysics Toolbox. *Spatial Vision, 10*(4), 433–436. <https://doi.org/10.1163/156856897X00357>

Braver, T. (2012). The variable nature of cognitive control: a dual mechanisms framework. *Trends In Cognitive Sciences*, *16*(2), 106-113. <https://doi.org/10.1016/j.tics.2011.12.010>

Conley, M., Dellarco, D., Rubien-Thomas, E., Cohen, A., Cervera, A., Tottenham, N., & Casey, B. (2018). The racially diverse affective expression (RADIATE) face stimulus set. *Psychiatry Research, 270*, 1059-1067. <https://doi.org/10.1016/j.psychres.2018.04.066>

Egner, T., & Hirsch, J. (2005). Cognitive control mechanisms resolve conflict through cortical amplification of task-relevant information. *Nature Neuroscience*, *8*(12), 1784–1790. <https://doi.org/10.1038/nn1594>

Eriksen, B. A., & Eriksen, C. W. (1974). Effects of noise letters upon the identification of a target letter in a nonsearch task. *Perception & Psychophysics, 16*, 143–149. <https://doi.org/10.3758/BF03203267>

Gratton, G., Coles, M., & Donchin, E. (1992). Optimizing the use of information: Strategic control of activation of responses. *Journal Of Experimental Psychology: General*, *121*(4), 480-506. <https://doi.org/10.1037/0096-3445.121.4.480>

Harvey, L. O. (1992). The critical operating characteristic and the evaluation of expert judgment. *Organizational Behavior and Human Decision Processes*, *53*(2), 229-251. <https://doi.org/10.1016/0749-5978(92)90063-d>

Kirchner, W. K. (1958). Age differences in short-term retention of rapidly changing information. *Journal of Experimental Psychology, 55*(4), 352–358. <https://doi.org/10.1037/h0043688>

Lafayette Instrument (2023). Model 32020A Purdue Pegboard Manual. <https://lafayetteinstrument.com/downloads/manuals/MAN-32020A-pdf-rev3.pdf> Accessed 14 Jan 2023.

Lundqvist, D., Flykt, A., & Öhman, A. (1998). The Karolinska directed emotional faces (KDEF). *CD ROM from Department of Clinical Neuroscience, Psychology section, Karolinska Institutet*, *91*(630), 2-2.

Miller, G. A. (1956). The magical number seven, plus or minus two: Some limits on our capacity for processing information. *Psychological Review, 63*(2), 81–97. <https://doi.org/10.1037/h0043158>

Stroop, J. R. (1935). Studies of interference in serial verbal reactions. *Journal of Experimental Psychology, 18*(6), 643–662. <https://doi.org/10.1037/h0054651>

The MathWorks Inc. (2022). Matlab version: 9.12 (R2022a), Natick, Massachusetts: The MathWorks Inc. <https://www.mathworks.com>
